# Development and Validation of ‘Patient Optimizer’ (POP) Algorithms for Predicting Surgical Risk with Machine Learning

**DOI:** 10.1101/2022.10.03.22280539

**Authors:** Gideon Kowadlo, Yoel Mittelberg, Milad Ghomlaghi, Daniel Stiglitz, Kartik Kishore, Ranjan Guha, Justin Nazareth, Laurence Weinberg

## Abstract

**Background:** Pre-operative risk assessment can help clinicians prepare patients for surgery, reducing the risk of perioperative complications, length of hospital stay, readmission and mortality. Further, it can facilitate collaborative decision-making and operational planning.

**Objective:** To develop effective pre-operative risk assessment algorithms (referred to as Patient Optimizer or POP) using Machine Learning (ML) that predicts the development of post-operative complications and provides pilot data to inform the design of a larger prospective study.

**Methods:** After institutional ethics approval, we developed a baseline model that encapsulates the standard manual approach of combining patient-risk and procedure-risk. In an automated process, additional variables were included and tested with 10-fold cross-validation, and the best performing features were selected. The models were evaluated and confidence intervals calculated using bootstrapping. Clinical expertise was used to restrict the cardinality of categorical variables (e.g. pathology results) by including the most clinically relevant values. The models were created with extreme gradient-boosted trees using XGBoost [1]. We evaluated performance using the area under the receiver operating characteristic curve (AUROC) and the area under the precision-recall curve (AUPRC). Data was obtained from a metropolitan university teaching hospital from January 2015 to July 2020. Data collection was restricted to adult patients undergoing elective surgery.

**Results:** A total of 11,475 adult admissions were included. For predicting the risk of any postoperative complication, kidney failure and length-of-stay (LOS), POP achieved an AUROC (95%CI) of 0.755 (0.744, 0.767), 0.869 (0.846, 0.891) and 0.841 (0.833, 0.847) respectively and AUPRC of 0.651 (0.632, 0.669), 0.326 (0.293, 0.359) and 0.741 (0.729, 0.753) respectively. For 30-day readmission and in-patient mortality, POP achieved an AUROC (95%CI) of 0.61 (0.587, 0.635) and 0.866 (0.777, 0.943) respectively and AUPRC of 0.116 (0.104, 0.132) and 0.031 (0.015, 0.072) respectively.

**Conclusion:** The POP algorithms effectively predicted any post-operative complications, kidney failure and LOS in the sample population. A larger study is justified to improve the algorithm to better predict complications and length of hospital stay. A larger dataset may also improve the prediction of additional specific complications, readmissions and mortality.

## 1 Introduction

The adoption and deployment of electronic health records (EHRs) has facilitated the accessibility of large patient datasets. Machine learning (ML) has succeeded in diverse arenas, demonstrating an ability to operate on large and complex datasets. At the intersection of EHR data and the progress of ML, is an opportunity to develop tools for personalised medicine. Currently, the most common ML applications in medicine are in imaging [2, 3]. An upcoming frontier is surgical risk prediction [4].

Surgery is often the only option to alleviate disability and reduce the risk of death from common conditions. Millions of people annually undergo surgical treatment, and surgical interventions account for an estimated 13% of the world’s total disability-adjusted life years (DALYs). Even in the most advanced hospital systems, there is a high mortality and complication rate [5, 6], risks of direct harm to patients and high financial costs. The WHO recognises these issues as major worldwide health burdens [7]. Fortunately, up to 50% of these complications are preventable [8].

Pre-operative risk assessment allows clinicians to mitigate adverse outcomes, better inform patients and their families about surgical outcomes and risks and plan post-operative care [9, 10]. The first generation of risk calculators exists, such as the American College of Surgeons National Surgical Quality Improvement Program (NSQIP) [11] and the Surgical Outcome Risk Tool (SORT) [12]. They are based on linear statistical techniques and are designed to use a low number of input parameters to be convenient for manual data entry. These approaches do not exploit the data available in modern EHR systems. Additionally, most provide mortality risk only. There are also manual risk assessments such as American Society of Anesthesiologists (ASA) Physical Status Classification [13] that are effective but subjective. It is often difficult for clinicians to find the data and calculate the score manually; therefore, they are rarely used [14].

In recent years, more sophisticated algorithms have been developed using ML. They typically predict a wider range of outcomes than traditional risk calculators and incorporate a larger set of input features made available by EHR data. The most common prediction outcomes are mortality and complications. ML can be more effective than traditional methods [14] such as ASA, CCI, POSSUM [15] and NSQIP [16] and can be more effective than human experts [17]. Various techniques have been used such as deep learning [18, 19], logistic regression [20, 21], generalised additive models (GAMs) [6] and decision trees [22, 23, 24, 16]. Further, some studies focus on harmonising EHR data [18], testing existing approaches on suitability for local populations [4, 20] or predicting the use of the readmission prevention clinic [22].

Most of the studies in the literature cited above, focus on the prediction of mortality and complications; however, additional endpoints are clinically meaningful. Some studies such as [18], utilise sequences of vital sign measurements, unstructured notes and radiological images, but in many hospitals, those data are not practically obtainable.

### 1.1 Study aims

This study aims to use readily available EHR data to develop interpretable ML risk prediction algorithms to standardise and improve clinical decision-making. The target endpoints are length-of-stay (LOS), complications, 30-day readmission and in-patient mortality. Our definition of readily available EHR data is patient history excluding unstructured notes and radiological imaging. The algorithms should be interpretable as the ultimate objective is to provide information that is understandable, actionable and trusted in a clinical setting.

## 2 Method

### 2.1 Study design

This was a single-centre cohort study with retrospective data collection in adult patients (aged ≥ 18 years) who underwent any elective surgical procedure at Austin Health between 1st January 2015 and 31st July 2020. Austin Health is a university teaching hospital in Australia, with a high volume of surgeries across multiple sub-specialities that are performed annually. We restricted cases to elective surgery where there is the greatest opportunity to mitigate risk.

We developed risk assessment models for the target endpoints following the Transparent Reporting of a Multivariable Prediction Model for Individual Prognosis or Diagnosis (TRIPOD) guidelines for risk prediction [25].

Our approach was to begin with a baseline model that emulates a standard approach internationally for surgical risk assessment, exploiting two key dimensions: patient-risk and procedure-risk. Each was derived from clinical expertise provided by perioperative clinicians with at least 10-years of postgraduate experience and familiarity with risk stratification for surgical morbidity. The next step was to iteratively add features to the baseline model, resulting in a unique set of features for each model.

### 2.2 Model development

Accounting for evidence indicating that ML can outperform standard risk predictors such as ASA, CCI and NSQIP and that simpler linear models are not as effective [14, 15, 16] we selected one ML architecture. The ML architecture that we selected was extreme gradient-boosted decision trees, using the XGBoost package [1], which are interpretable and among the best performing for tabular health data.

There are four main stages to the method. The Data source provides data for Pre-processing that reduces dimensionality and transforms relational data into a tabular form suitable for algorithm consumption. Feature selection selects a subset of features to optimise prediction scores for each endpoint. Finally, an Evaluation of the model is performed with bootstrapping. The pipeline is illustrated in Figure 1 and elaborated below.

**Figure 1:**
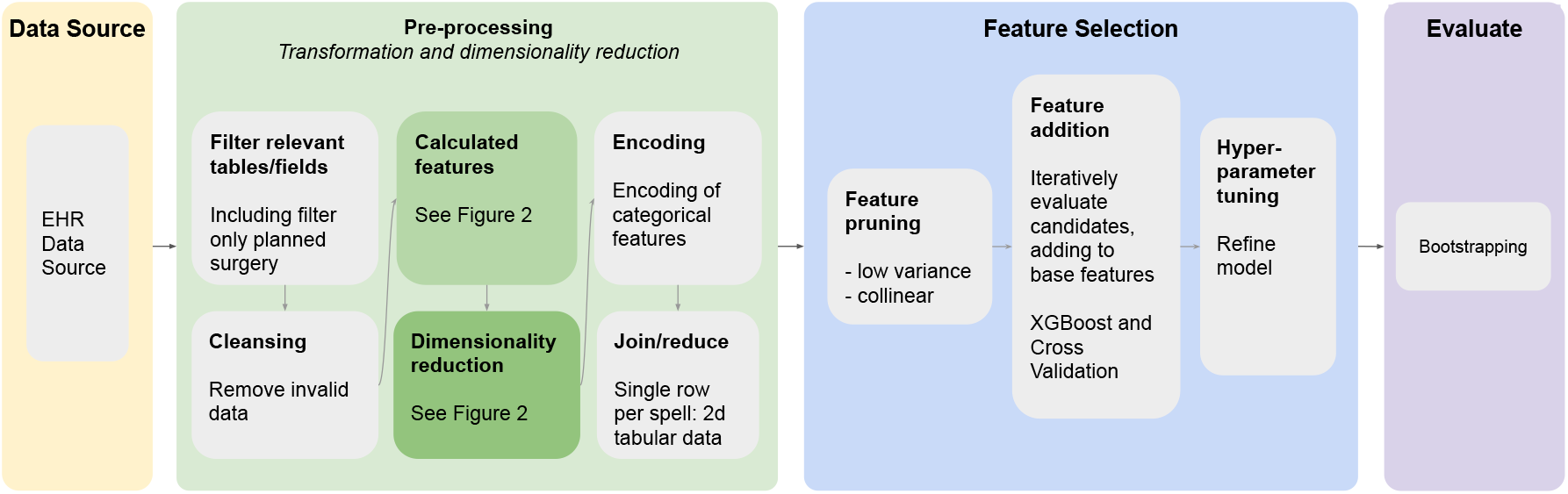
Data processing pipeline.

#### 2.2.1 Data source

The Data Analytics and Research Evaluation (DARE) Centre provided a data extract from the Austin Health Cerner EHR system. A total of 11,475 unique admissions were included, covering all elective adult surgical procedures.

The raw data includes:

- patient demographic details (age, weight, height, gender)
- procedures performed (primary/scheduled and other)
- other procedural information (including details of the admission and episode)
- pathology results
- medications prescribed during admission
- comorbidities: diagnoses using the International Classification of Diseases (ICD-10-AM, 9th Edition)
- Charlson comorbidity index (CCI) derived from the ICD codes
- complications, indicated by ICD codes

#### 2.2.2 Pre-processing

After cleansing the data, we derived features from raw values: body mass index (BMI) and the two features used for the baseline model, namely estimates of patient-risk and procedure-risk (Section 2.1). The process is illustrated in Figure 2.

**Figure 2:**
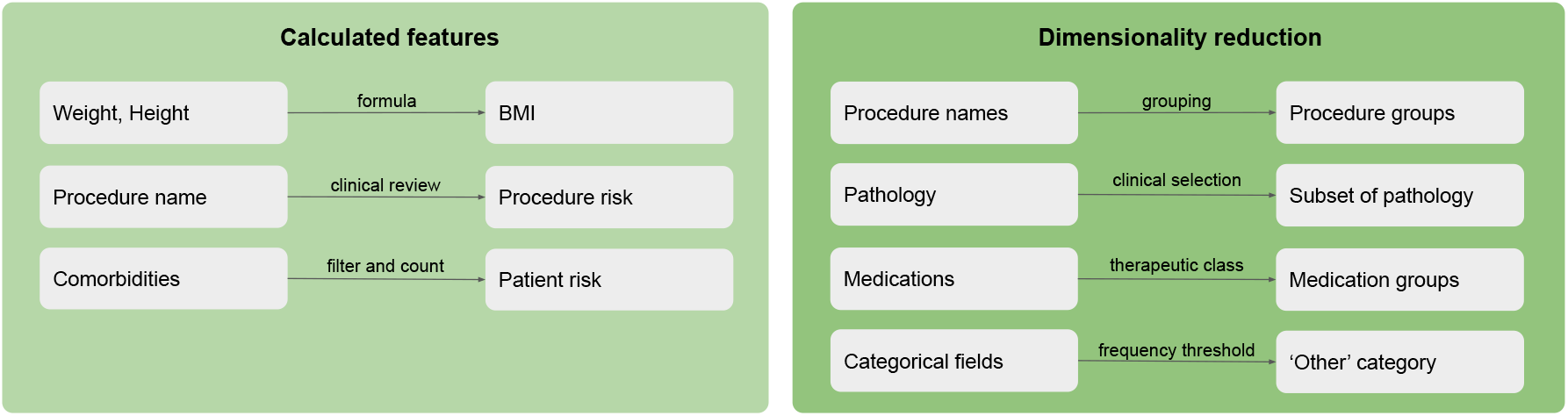
Dimensionality reduction and calculated features.

Patient-risk is a proxy for ASA [13]. It is an ordinal numerical value calculated through the number of patient diagnoses (using ICD codes), with an additional increment for ‘cancerous’ and ‘cardiopulmonary and vascular’ conditions. Procedure-risk is an ordinal categorical value calculated using a clinically determined risk rating (low, medium or high) of the scheduled surgical procedure, which was estimated as the earliest non-preparatory procedure.

We reduced dimensionality where possible to reduce overfitting and improve interpretability. We grouped procedure names by anatomical region. Although all patient episodes are elective, the individual procedures can be varying levels of elective or emergency, referred to as procedure type. The procedure type was reduced to a binary category. For laboratory results, we selected a priori eight clinically relevant variables, namely haemoglobin, albumin, creatinine, urea, international normalised ration, platelet count, activated partial thromboplastin time, and estimated glomerular filtration rate. We grouped patient medications by therapeutic class. Finally, very infrequent categories were grouped into an ‘other’ bucket. Dimensionality reduction is summarised in Figure 2.

Categorical data was one-hot encoded. Where there were one-to-many relationships (such as admission to medications) the reduction methods were chosen to provide the most clinically relevant summary. As XGBoost is a decision tree-based model, standardisation of numerical features was not necessary.

Missing data were treated as legitimate input, either by creating a ‘missing’ category or utilising XGBoost’s in-built mechanism.

See Appendix 5.1 for more details on pre-processing.

#### 2.2.3 Feature selection

Highly correlated (or collinear) features were removed due to their redundancy. We used the variance inflation factor method for multi-collinearity analysis with a threshold of 10 [26, 27]. Variables with very low variance were removed by detecting features where the ratio between the highest occurring value and the second highest was greater than 19, a large threshold to avoid losing valuable information [28].

After training and scoring the baseline model consisting of patient-risk and procedure-risk (Section 2.1), an automated iterative process added and tested new features. Each available feature was individually added to the model and evaluated using area under the receiver operating characteristic curve (AUROC) with 10-fold cross-validation. The feature that achieved the highest gain in score was added to the selected feature set and the search restarted. The remaining features were re-tested in subsequent iterations, after which the composition of the selected feature set had changed. The process continued until all features were used and then the model with the highest score was selected.

Hyperparameter tuning then took place to optimise results (see Appendix 5.2).

### 2.3 Predicted outcomes

Length-of-stay (LOS) was framed as a multiclass classification. We identified three dominant groupings through visual inspection of the distribution (see Figure 3) and defined them by ordering and then segmenting the data into three equally sized buckets. The resulting groups were low (≤31 hours), medium (31 – 117 hours), and high (≥117 hours), equating to one night, two to four nights and five nights or more. The ranges were validated through clinical review. There was a classifier for each bucket and the prediction was the classifier with the highest confidence.

**Figure 3:**
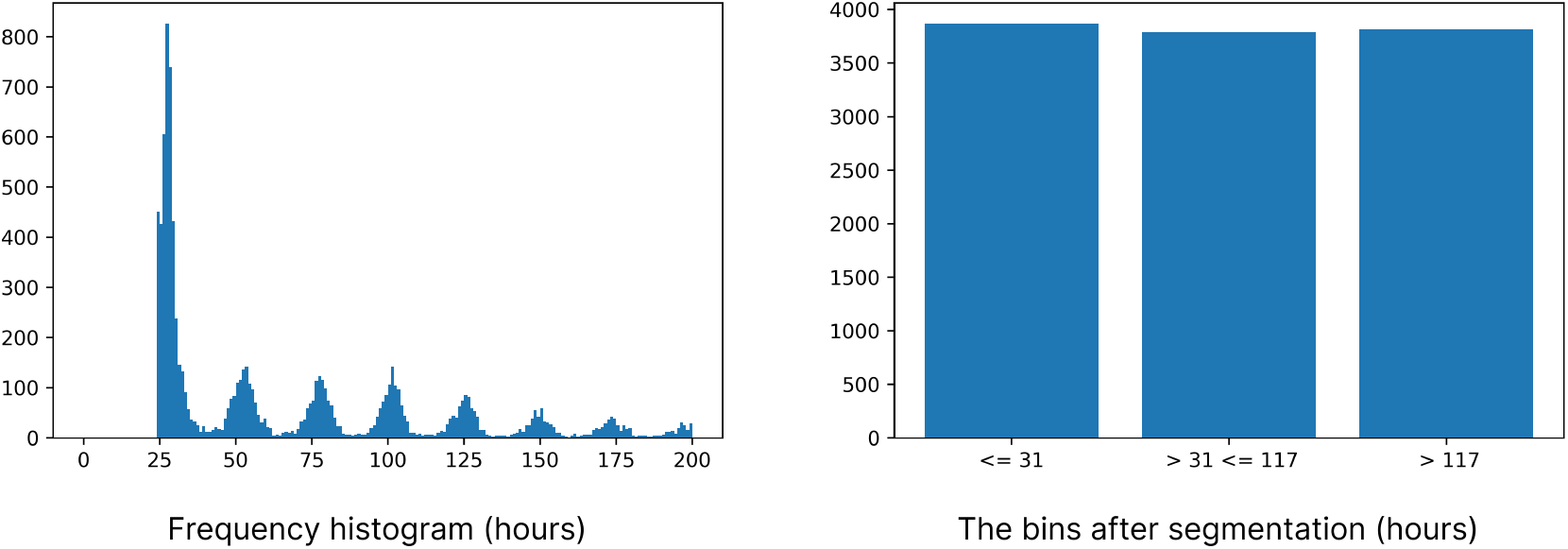
Length-of-stay: Training data are segmented into 3 classes, to cast predicting length-of-stay as multiclass classification. There is a clear periodicity around whole days. The x-axis is truncated at 200 hours to provide detail in the most interesting range. The trend continues past 200 hours with a steady monotonic decrease in magnitude.

Unplanned 30-day readmission, in-patient mortality and the presence of complications (as indicated by the ICD code) were explicitly labelled in the dataset.

### 2.4 Evaluation

The final score and confidence intervals were calculated with non-parametric bootstrapping using 1,000 iterations. For each iteration the training set size was the same as the whole dataset. As bootstrapping involves sampling with replacement, this resulted in approximately 70% unique samples for training, leaving the left-out 30% for testing.

#### 2.4.1 Performance metrics

Several metrics were used to assess and measure performance: area under the receiver operating characteristic curve (AUROC), area under the precision-recall curve (AUPRC) and F1 (FBeta, where beta = 1). AUROC is most common in related literature. A drawback of AUROC is that it can be misleading on extremely rare classes such as mortality and readmission. In such cases, it can achieve an artificially high score because the true negatives dwarf the false positives^1^. AUPRC is more informative with extremely rare labels [29, 15]. It indicates the trade-off between precision (the proportion of true positives of all predicted positives), also referred to as positive predictive value (PPV), and recall (the proportion of true positives of all positives). F1, the harmonic mean of precision and recall, is also suitable for rare classes. We used micro-averaging to calculate the area under the curve for multiclass predictions.

In addition to the single metric derived from the ‘area’ under the respective curves AUROC and AUPRC, we also inspected the profiles of the curves, showing how they perform at different operating points.

#### 2.4.2 Interpretability

To visualise the relative importance of features for each model, we utilised two methods. The first was XGBoost feature importance, based on the average gain of splits per feature. The other was SHapley Additive exPlanations (SHAP) [30] which uses cooperative game theory to assign partial credit to the input variables for the model’s output. Both methods indicate feature importance from different perspectives. XGBoost feature importance gives direct insight into the internal structure of the learned trees and provides a single absolute value for importance. SHAP treats the model as a black box and bases the importance on the observed behaviour of the model. The plots are more informative, showing the distribution of observed values and the corresponding directionality of the impact on the model.

To visualise the features’ influence on specific predictions for individual patients, we used SHAP. We plotted typical true positives for each of the effective models to demonstrate how SHAP can be used to help make specific predictions actionable.

## 3 Results

### 3.1 Data characteristics

A total of 11,475 adults were included. There were 41 (0.36%) occurrences of in-patient mortality and 941 (8.2%) occurrences of 30-day readmissions. There were 4,351 (37.92%) complications. The number of occurrences of low, medium and high LOS were 3,868 (33.7%), 3,790 (33.0%) and 3,817 (33.3%), respectively. the data characteristics are presented in Table 1.

**Table 1:** **Data characteristics:** The first column shows the number of affirmative cases for binary fields and the number of unique values for multivalue categorical fields. The second column shows the number of admissions with a valid value (e.g., if height is missing, it is deemed invalid). Empty cells denote N/A.

|  | # Affirmative /<br># Categories | # Valid values | Median | Mean (SD) |
| --- | --- | --- | --- | --- |
| **Demographics** |  |  |  |  |
| M | 6,234 (54.33%) | 11,475 (100.00%) |  |  |
| F | 5,241 (45.67%) | 11,475 (100.00%) |  |  |
| Age |  | 11,475 (100.00%) | 62.00 | 59.24 (17.81) |
| Height |  | 4,958 (43.21%) | 167.00 | 166.01 (14.63) |
| Weight |  | 6,920 (60.31%) | 81.00 | 84.13 (21.46) |
| **Derived features** |  |  |  |  |
| BMI |  | 4,931 (42.97%) | 29.38 | 32.10 (14.09) |
| Procedure-risk |  | 11,475 (100.00%) | 1.00 | 1.63 (0.71) |
| Patient-risk |  | 11,475 (100.00%) | 3.00 | 4.06 (3.13) |
| **Other** |  |  |  |  |
| Emergency procedure | 1,416 (12.34%) | 11,475 (100.00%) |  |  |
| **Categorical fields** |  |  |  |  |
| Procedures | 911 |  |  |  |
| Medication | 923 |  |  |  |
| Pathology | 75 |  |  |  |

### 3.2 Accuracy

The results are summarised in Tables 2 and 3, and the ROC and PR curves are shown in Figures 4 and 5. We selected only those specific complications with an adequate number of positive examples to make training feasible (above a threshold of 100 (0.8%)).

**Table 2:** Performance of risk models

| Prediction | # Cases<br>(prevalence) | AUROC (95% CI) | AUPRC (95% CI) | F1 Value (95% CI) |
| --- | --- | --- | --- | --- |
| In-patient mortality | 41 (0.36%) | 0.866 (0.777, 0.943) | 0.031 (0.015, 0.072) | 0.057 (0.000, 0.167) |
| 30-day re-admission | 941 (8.20%) | 0.610 (0.587, 0.635) | 0.116 (0.104, 0.132) | 0.122 (0.078, 0.156) |
| Length-of-stay | N/A | 0.841 (0.833, 0.847) | 0.741 (0.729, 0.753) | 0.666 (0.654, 0.678) |

**Table 3:** Performance of risk models for complications

| Prediction | # Cases<br>(prevalence) | AUROC (95%<br>CI) | AUPRC (95%<br>CI) | F1 Value (95%<br>CI) |
| --- | --- | --- | --- | --- |
| Any complication | 4,351<br>(37.92%) | 0.755 (0.744, 0.767) | 0.651 (0.632, 0.669) | 0.621 (0.602, 0.639) |
| Heart failure | 116 (1.01%) | 0.835 (0.773, 0.887) | 0.101 (0.055, 0.181) | 0.141 (0.097, 0.19) |
| Delirium | 303 (2.64%) | 0.827 (0.793, 0.857) | 0.139 (0.099, 0.187) | 0.189 (0.153, 0.225) |
| Arrhythmia | 341 (2.97%) | 0.794 (0.764, 0.822) | 0.122 (0.092, 0.165) | 0.148 (0.129, 0.169) |
| Kidney failure | 505 (4.40%) | 0.869 (0.846, 0.891) | 0.336 (0.282, 0.390) | 0.326 (0.293, 0.359) |

**Figure 4:**
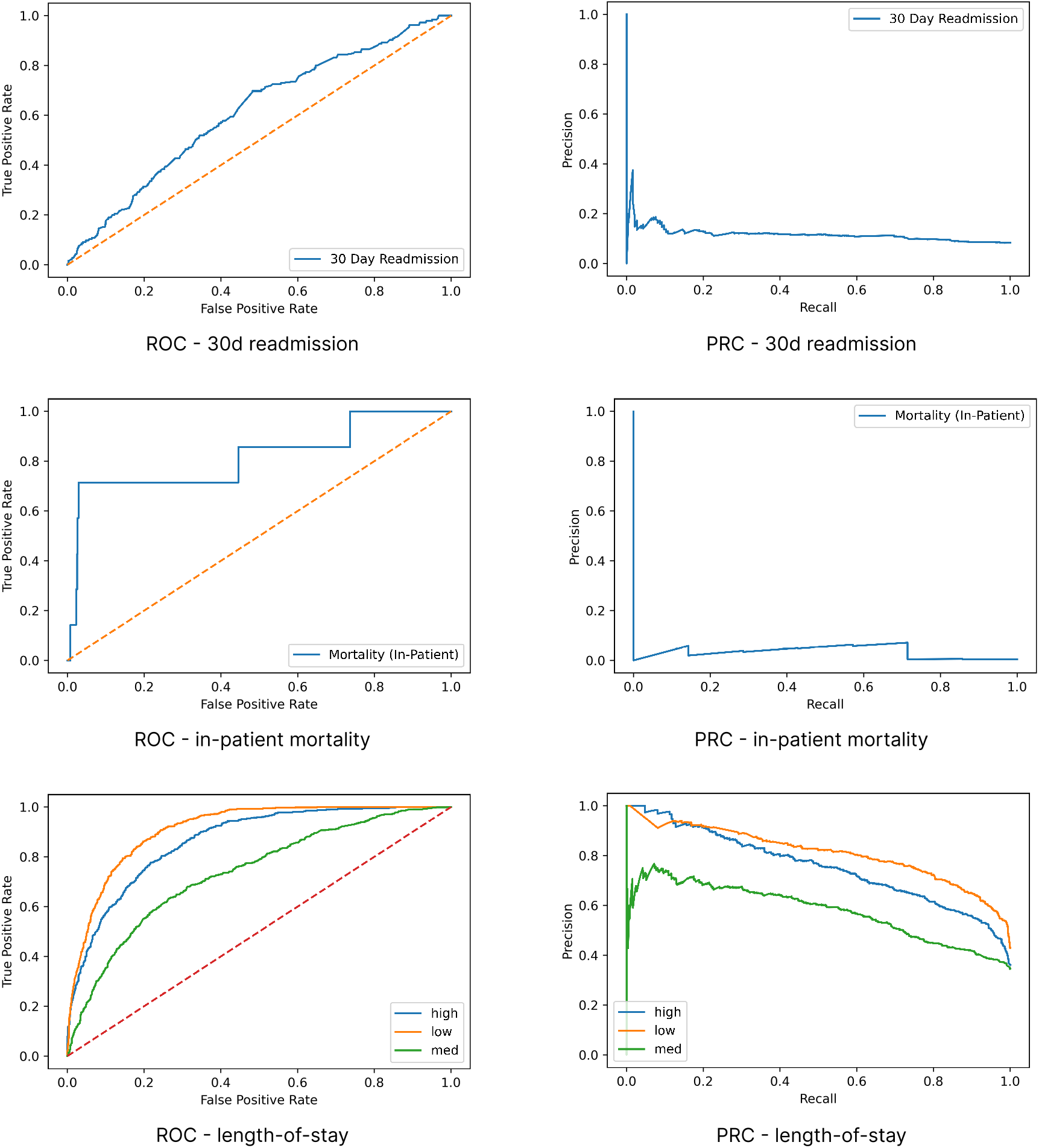
Receiver operating characteristic and precision-recall curves – readmission, mortality and length-of-stay.

**Figure 5:**
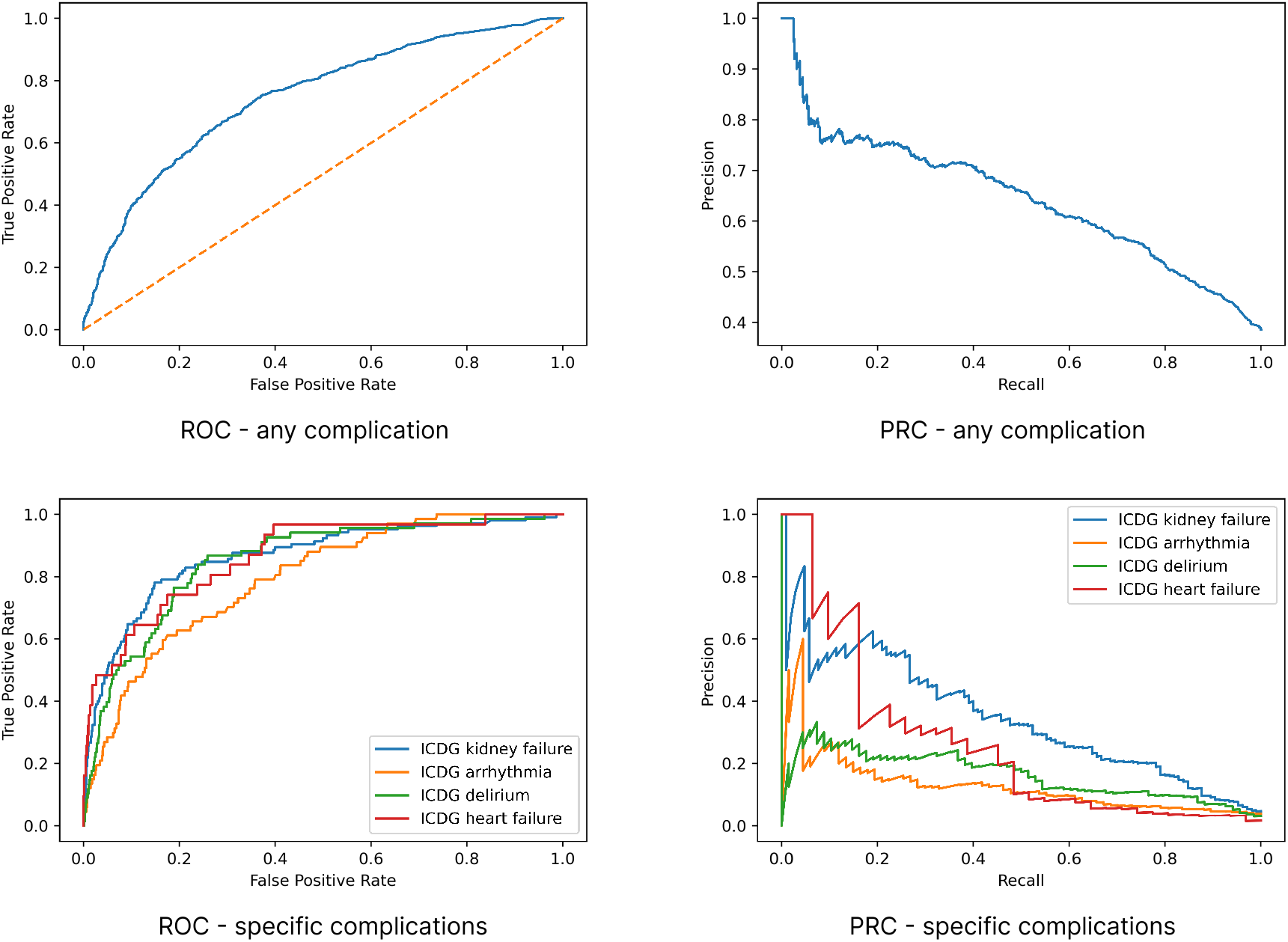
Receiver operating characteristic and precision-recall curves – complications.

For predicting the risk of any post-operative complication, kidney failure and LOS, POP achieved an AUROC (95%CI) of 0.755 (0.744, 0.767), 0.869 (0.846, 0.891) and 0.841 (0.833, 0.847) respectively and AUPRC of 0.651 (0.632, 0.669), 0.336 (0.282, 0.390) and 0.741 (0.729, 0.753), respectively. Refer to table for full results of other specific complications. For 30-day readmission and in-patient mortality, POP achieved an AUROC (95%CI) of 0.61 (0.587, 0.635) and 0.866 (0.777, 0.943), respectively and AUPRC of 0.116 (0.104, 0.132) and 0.031 (0.015, 0.072), respectively.

On visual inspection, the ROC curves provide reasonable operating points for all models. Inspection of the precision-recall (PR) curves also shows some models have effective operating points. Still, the endpoints with extremely rare positive examples do not account for readmission, mortality, and the specific complications other than kidney failure. For LOS, accuracy was consistently higher for the two ends of the spectrum (low and high) compared to medium which experienced more class overlap than low or high.

### 3.3 Interpretability

The selected features and their importance are shown for the effective models: complications in Figure 6, kidney failure in Figure 7 and LOS in Figure 8. For terminology used in the figures, please refer to Table 4.

**Table 4:**
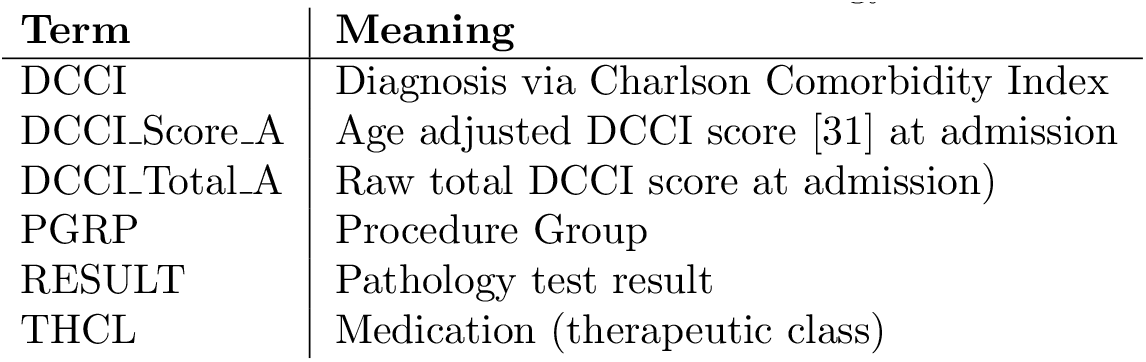
Feature-name terminology

**Figure 6:**
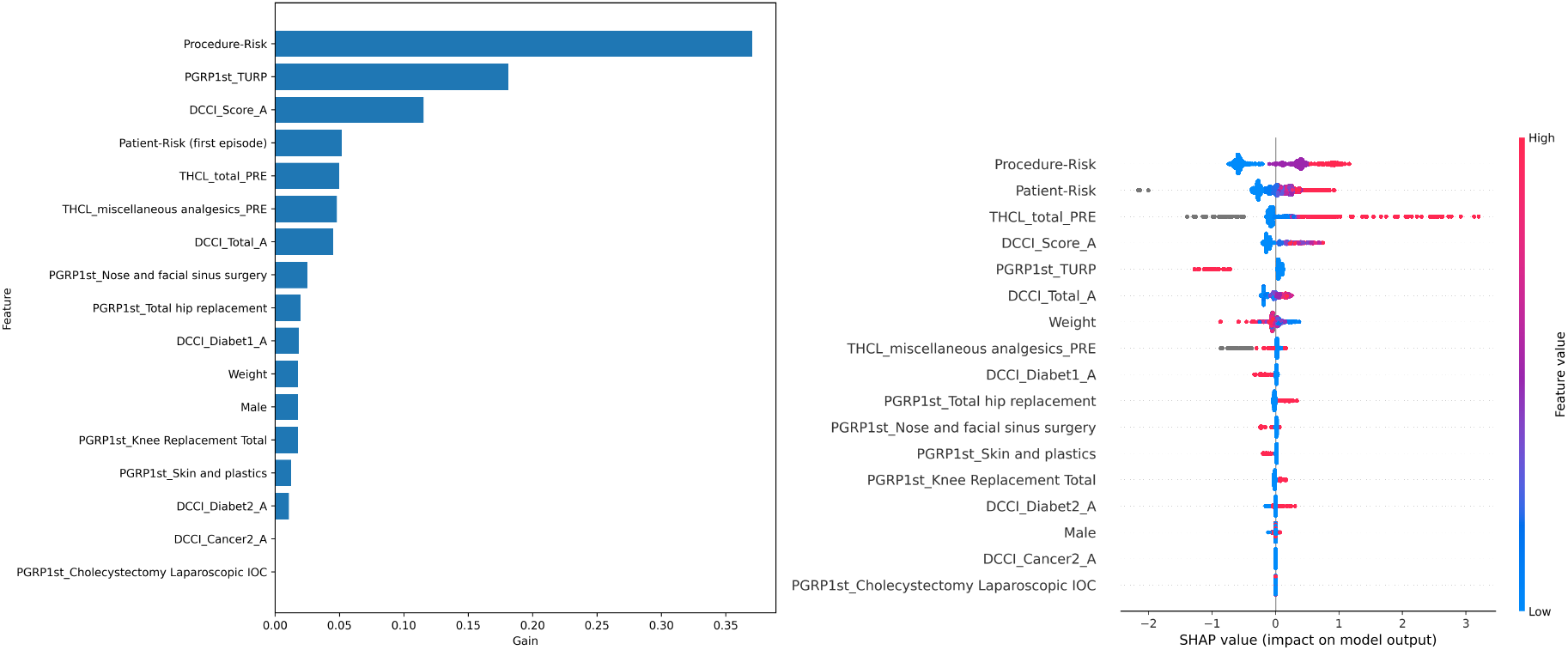
Feature importance for any complication: XGBoost gain (left) and SHAP (right), where each dot represents one sample, the colour indicates the value and the position on the x-axis indicates the impact (positive or negative) on model output. Refer to Table 4 for terminology.

**Figure 7:**
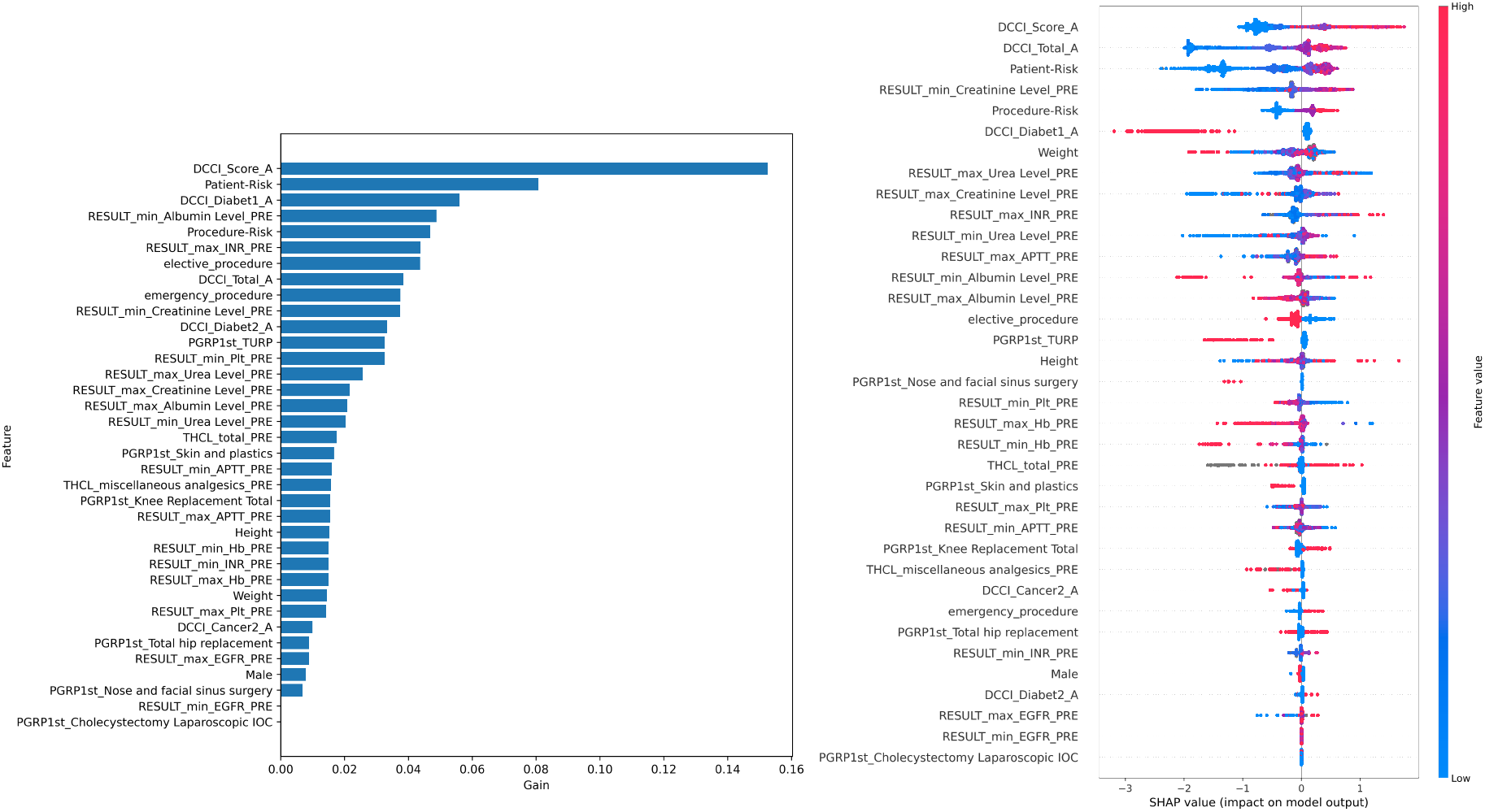
Feature importance for kidney failure: XGBoost gain (left) and SHAP (right), where each dot represents one sample, the colour indicates the value and the position on the x-axis indicates the impact (positive or negative) on model output. Refer to Table 4 for terminology.

**Figure 8:**
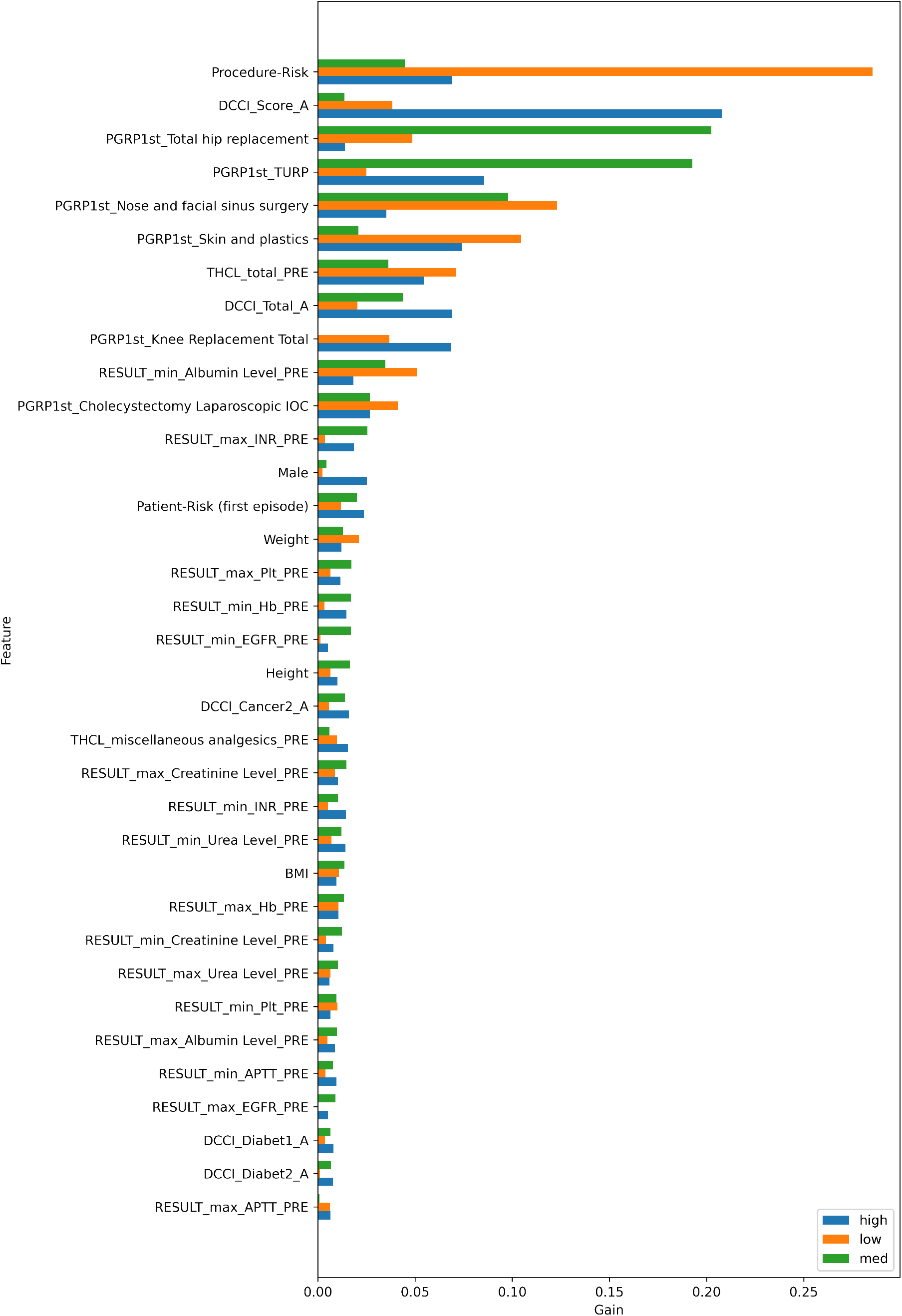
Feature importance for length-of-stay using XGBoost gain. Refer to Table 4 for terminology.

For all the models and visualisation methods, procedure-risk and features representing the patient’s health (CCI summaries and patient-risk) are amongst the top factors. Patient-risk and CCI represent the patient’s overall health. Although patient-risk is derived from more specific and diverse comorbidities than CCI, the feature importance plots showed that across the cohort, CCI was an important factor particularly in the age-adjusted CCI [31], and comparable to patient-risk. However, patient-risk and CCI are both valuable as they contain different information, as illustrated in the example of a specific patient high LOS, Figure 10, where patient-risk and CCI have an opposing influence.

In addition to procedure-risk and patient health, there are other important features. For any complication (Figure 6), XGBoost shows significant tree splits for some specific procedure groups: diabetes, total medication dosages and use of analgesics. The SHAP features are largely aligned, with differences in the relative values. For kidney failure (Figure 7), related morbidities (diabetes, cancer) and pathology results (albumin, creatinine, urea, activated partial thromboplastin time and haemoglobin) are also important. For length-of-stay (Figure 8), the features differing importance to the individual models (low, medium and high), although many features are unimportant for all models. Compared to the other models, specific procedure groups are relatively important. Feature importance in specific predictions using SHAP plots is shown for typical true positive cases of kidney failure in Figure 9 and a high LOS for a procedure with a medium-term LOS in Figure 10. The purpose is to show how SHAP can provide a convenient interpretation of the important factors for a given prediction.

**Figure 9:**
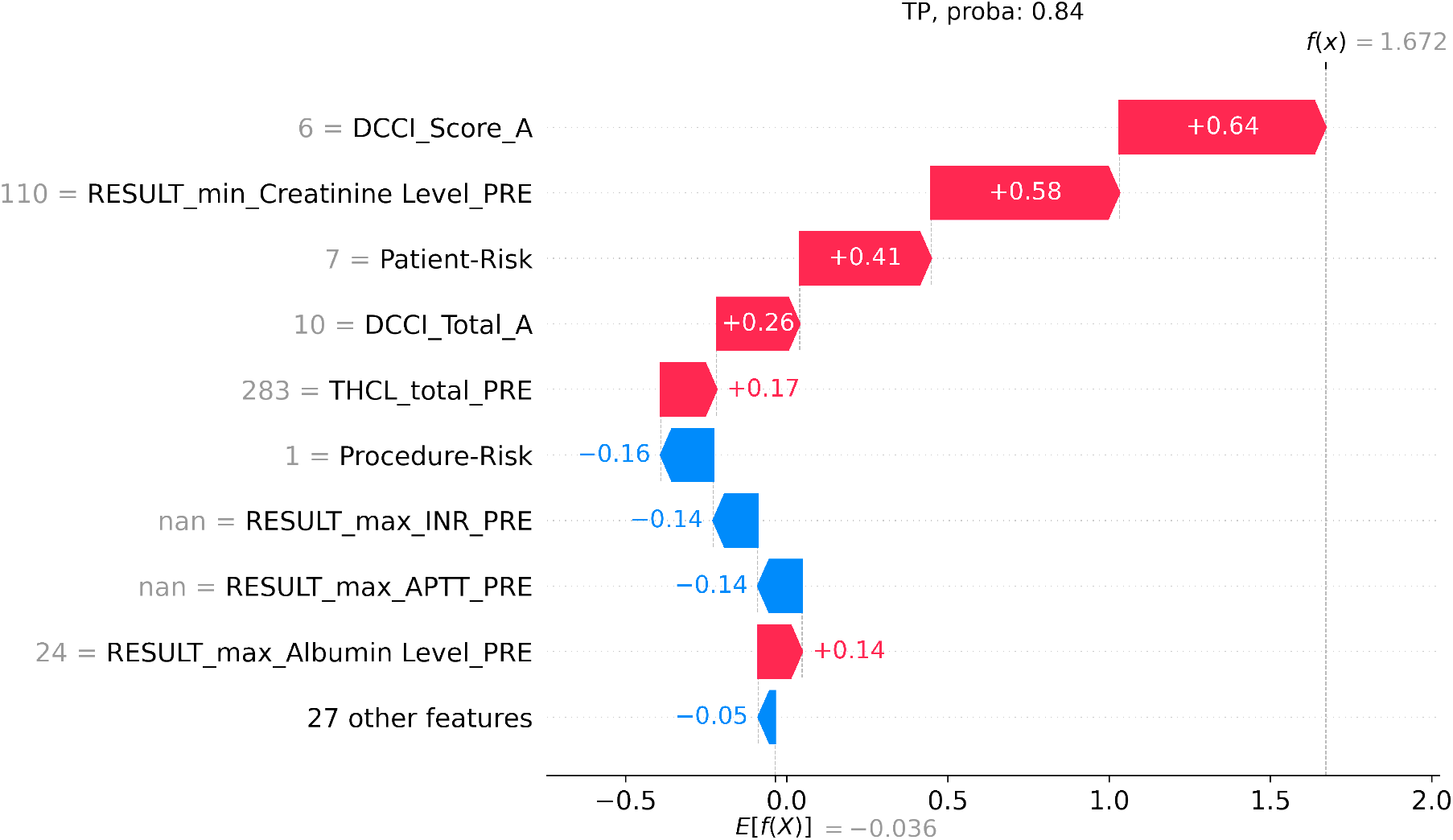
SHAP visualisation for a specific patient’s risk of a kidney failure. This is a True Positive (TP) prediction with a probability of 0.87. The length of the bar indicates the influence of that feature on the prediction. The colour indicates whether the influence is positive (red) or negative (blue). The grey value to the left of the feature name is the value of that feature for this patient.

**Figure 10:**
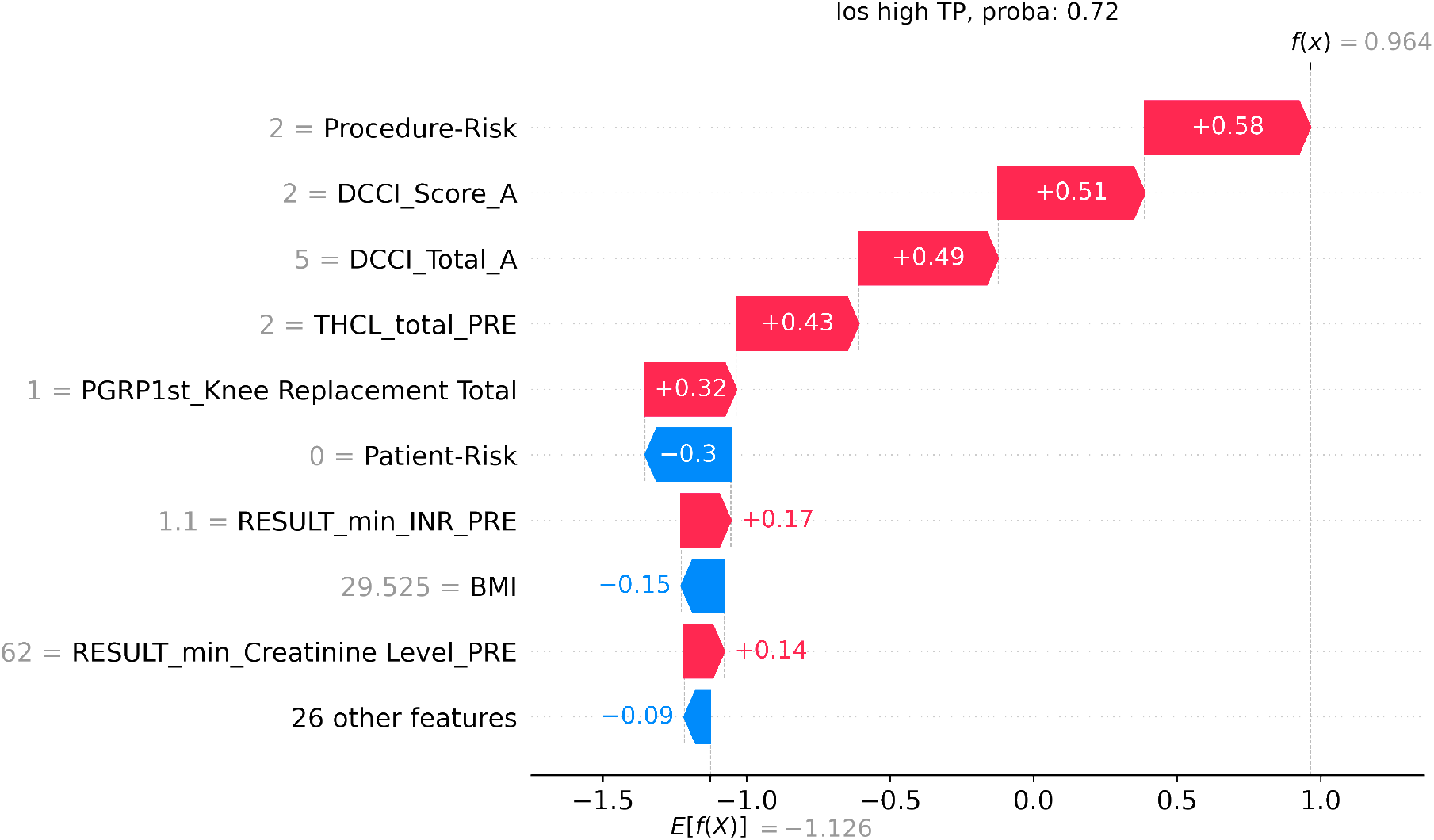
SHAP visualisation for a specific patient’s short length-of-stay. This is a True Positive (TP) prediction of a high length of stay, with a probability of 0.72. The length of the bar indicates the influence of that feature on the prediction. The colour indicates whether the influence is positive (red) or negative (blue). The grey value to the left of the feature name is the value of that feature for this patient.

## 4 Discussion

### 4.1 Key findings

In this single-centre cohort study in adult surgical patients, we developed effective pre-operative risk assessment algorithms (POP) using machine learning, providing pilot data to inform the design of a larger prospective study. We found that POP algorithms were effective for predicting post-operative complications and LOS. However, a larger study is justified to further improve the algorithm for predicting specific complications, readmission and mortality.

### 4.2 Comparison to other methods

Comparing accuracy to other models in the literature is very difficult for several reasons. The quality and structure of different datasets vary greatly, as cohort differences can influence results [32] and endpoints are often defined differently (e.g., 24 hours after admission compared to immediately before surgery). Moreover, the choice of performance metrics also varies. However, considering the difficulties, it can be useful to compare results to provide some context.

One of the studies that we compared to is [18]. Beyond tabular EHR data, they utilized additional data sources including radiological imaging, unstructured notes, vital sign measurements, time-series embedding to handle these data streams, as well as ensembling of complementary models. While we consider it to be the ‘gold-standard’, and therefore present it as context, we do not aim to equal their scores. Our study investigates the feasibility of risk predictions with more limited and commonly available data sets.

### 4.3 Evaluation metrics

The standard practice for evaluating risk prediction algorithms is the ROC curve. Using ROC, all of our models appear to be effective. They have a good profile with viable operating points, and relatively good AUROC. However the results using the PR curves reveal a different story. AUPRC for readmission and mortality is very low, and there are no satisfactory operating points on the profile. The results confirm that ROC can be misleading for rare classes as suggested by [15] (and discussed in Section 2.4). They demonstrate the importance of metrics that are insensitive to rare classes, such as AUPRC or FBeta for clinical algorithms. We used a relatively small dataset (see Section 4.7). With more data and therefore more positive examples, the performance is likely to improve, as measured by both AUROC and AUPRC.

### 4.4 LOS prediction

LOS classification was very effective. There is LOS data for every admission, providing ample training signal, which is reflected in the ROC and PR curves. LOS predictions have both clinical and operational benefits. From a clinical perspective, a ‘longer than expected’ stay prediction could prompt closer attention. From an operational perspective, these predictions could be used for scheduling to optimise for ward utilisation and selection of appropriate sites.

To the best of our knowledge, other ML risk predictors did not consider LOS, except [18]. They predicted ‘prolonged length of stay’, defined as ‘at least 7 days’, whereas POP predicts multivalue LOS: low, medium or high. Predicting multivalue LOS makes it possible to have a dynamic definition of ‘prolonged’ that depends on factors such as procedure and patient. For example, a medium stay (two to four nights) prediction could trigger ‘prolonged’ for short-stay surgery (1 night) and healthy patients. Secondly, a more granular prediction allows better operational planning. Our accuracy, measured using AUROC, was comparable to [18], 0.841 compared to 0.85 and 0.86 (for two hospital sites respectively), despite fewer data types and a much smaller dataset. Unfortunately AUPRC is unavailable for comparison to gain a fuller picture.

### 4.5 Complication prediction

Results for predicting any complication were promising, with both AUROC and AUPRC having viable operating points. The four specific complications with adequate positive examples to train a model had reasonable ROC curves, but precision and recall showed that only kidney failure, which is less rare than the others, was a viable model.

In a clinical setting, positive predictions could be used as a general indicator that a morbidity is possible, and investigations are warranted. An example of an operating point for kidney failure is approximately recall = 12%, precision = 62%. Out of 100 patients with kidney failure, the model will identify approximately 12. Of those, approximately 62% (7.4) of these patients will actually develop kidney failure (true positives). If the information is presented so that it doesn’t give a false sense of security if *not* shown, then it can pick up when there *is* a case, aiding clinical care.

The results compare favourably to similar studies, despite a much smaller dataset (Section 4.7). Across specific complications, and using AUROC, POP scored 0.798 – 0.869 compared to 0.820 – 0.940 [6], 0.772 – 0.909 [16] and 0.88 – 0.89 [18]. For any complication, POP scored 0.755 compared to 0.829 – 0.836 in [16]. Again, PR results are unavailable for a more complete comparison. Precision (referred to as PPV or positive predictive value) was reported in [6], which showed the same pattern as POP with rare classes (i.e., the rarer classes generally have lower precision).

### 4.6 Interpretability

The introduction of ML often leads to improved performance, but it can come at the cost of interpretability. We used XGBoost and SHAP feature importance plots. They are intuitive and build trust in the model, helping to make it understandable and actionable.

The relative importance of features learned by the algorithm aligns with clinical practice. For example, the high importance of procedure information combined with patient health is commonly used to assess the risk of surgery. Alignment with clinical practice provides confidence that learning is effective and generalisable. Additionally, the relative weighting of feature importances can provide new insights into the relationship between features and outcomes. Although not causative, it indicates a relationship, and warrants further investigation. A better understanding of the factors, especially modifiable ones, could impact clinical practice. The first type of visualisation is the feature importance of the model in general, indicating systematic relationships across samples in the dataset. The other type of visualisation was feature importance for specific predictions, which highlights factors for individual patients. This information can provide an opportunity for more personalised risk mitigation.

We now explore kidney failure as a case study. The model highlighted comorbidities (Figure 7) that align with current knowledge. It is also the likely explanation for a similarly protective effect in the model for any complication (Figure 6). Several pathology results are also considered important; for example, some known to be related to renal function such as creatinine and urea, and others that are generally indicative of post-operative outcomes such as albumin [5], pathology related to coagulation (INR, APTT, PLT) [33, 34] and heamaglobin (Hb) [35, 36]. Some procedure groups were protective: ‘trans-urethral resection of the prostate’ (TURP), likely because it improves renal function; and nose and ‘facial sinus surgery’, likely because it is very low risk. The importance of ‘total knee replacement’ is unexpected, and warrants further investigation; for example, the underlying cause may be tourniquet time, length of surgery or even anaesthetic type.

Surprisingly, diabetes is protective. We hypothesise that patients with this conditions are more actively managed, so it is not picked up by the model, which learns from raw correlations. A more thorough investigation that includes causal analysis is an important topic for future work.

Understanding the expected and unexpected features may allow for patient-specific pre-operative intervention to minimise post-operative complications. For example, by optimising HbA1c in diabetics, being aggressive in comorbid management such as blood pressure optimisation, and shortening tourniquet time in knee replacements.

It is possible that the patient with kidney failure (Figure 9) could have been missed, because they do not have diabetes and it was a low-risk procedure. However, the patient suffered post-operative kidney failure and POP predicted it with 84% confidence. High creatinine and comorbidity burden are the most significant contributions. The high creatinine confirm that this patient likely has impaired renal function, and the prediction could lead to pre-surgical intervention including more intensive management of medications, ensuring the patient is well hydrated, selecting more appropriate anaesthesia type, and optimisation and monitoring of renal perfusion.

Another case study is LOS. The patient underwent a knee-replacement procedure, which is usually a medium LOS. However, POP identified this patient as having a high LOS (above five nights) and the SHAP plot (Figure 10) provides visibility into the reasons. The most significant indicators are comorbidities, a high number of prescribed medications and the procedure itself. As a result of the prediction, and seeing the concrete reasons, the patient could be booked in for a longer stay. Additionally, identification of knee replacement is more impetus to investigate the process of knee replacements, as discussed for kidney failure above.

Most of the studies reviewed, consider interpretability of models to be important for clinical practice, chose algorithms that support it [16] and additionally investigated and reported interpretability results [14, 22, 6, 37, 15, 38]. Lee et al. [38] used a GAM-NN for the benefit of neural networks and the interpretability of GAMs–there is a neural network for each input feature (or group of features), and they are linearly combined for classification.

However, most studies did not consider which features contributed to specific predictions. Although Bihorac et al. [6] used an approach, where the feature importance was “based on how different she or he is from the patient with an ‘average’ risk”. The reason for the prediction must be inferred indirectly, but the method could be applied to any model. Rajkomar et al. [18] used deep learning neural networks, where interpretability is more of a challenge. They showed a proof-of-concept of how it can be done. Active research is taking place to improve interpretability of deep learning models [39]. SHAP plots that were demonstrated here, can be used with any model.

### 4.7 Limitations

The dataset is relatively small for this type of algorithm. For context, other studies cited in this paper range between 51,457 patients [17] to 99,755 [16] admissions and [18] 216,221 admission. We expect the performance to improve with more data, particularly for specific complications, readmission and mortality, as there were very few positive examples from which to learn in our study.

The booked procedure is an important factor for predicting risk, according to both clinical practice and the models’ feature importance. However, the booked procedure was not explicitly labelled and was therefore estimated (see Section 2.2.2), resulting in errors that were difficult to quantify.

Data for patient height and weight were sparse, but these fields are considered to be important patient health factors. Likewise, there were many cases of missing medication therapeutic class, leading to information loss when grouping medications by this variable. Obtaining additional data in these respects is likely to improve performance.

The dataset did not extend beyond discharge, restricting mortality to in-patient mortality. In comparison, most risk calculators predict mortality at various stages after discharge such as 30-day and 60-day mortality. This is clinically important and there would be more examples which would improve the model.

### 4.8 Future work

In future, well-known applied ML techniques for medical risk prediction could be used to improve the initial results; for example, class balancing and model ensembling [40] and data augmentation [19]. There is also scope to explore alternative feature engineering, such as using additional derived features regarding previous admissions, other encoding methods for categorical variables, learning a lower dimensional space for categorical features using decision trees [6], and including additional categories for tests and medications that were ignored in this study.

Another major area of interest is continual risk assessment throughout the admission, including in the post-operative period up until discharge. Only a few related studies considered risk assessment after surgery [38, 18, 22]. It is important because decisions are made throughout the admission and post-surgical care also has the potential to help avoid complications, readmission and mortality.

## 5 Conclusions

In this study, we developed novel algorithms (POP) that exploit tabular EHR data to predict surgical patient outcomes. The algorithms were effective for post-operative complications and LOS in this patient population, but ineffective for predicting readmission and mortality due to extremely rare cases. The results reinforce the importance of using metrics that are suitable for rare cases, which is uncommon in other surgical risk prediction studies. A larger study is justified to improve the algorithms in better predicting complications and length of hospital stay. A larger dataset may also improve the prediction of readmissions and mortality, which were extremely rare. Together with interpretable feature importance plots, surgical risk predictions provide clinically relevant information, that may help to mitigate risks and improve patient outcomes.

## Data Availability

The datasets generated and/or analysed during the current study are not publicly available as there was no data sharing as part of the ethics approval (and raw data is potentially re-identifiable) but are available from the corresponding author on reasonable request.

## Acknowledgements

Thanks to David Gyorki and Dave Rawlinson for valuable discussions and advice at every stage.

## Ethics approval and consent to participate

A waiver of ethical approval was received from Austin Health Office for Research (approval number: 38679; approval date: 23rd March 2020); written informed consent was waived because de-identified retrospective data were used.

## Consent for publication

NA

## Authors’ contributions

GK designed the study and drafted the manuscript. YM helped to design the methodology and led the implementation and analysis. MG contributed to implementation and analysis. KK extracted the data and assisted with data interpretation and analysis. DS helped to design the study from a clinical perspective. DS, RG, JN and LW provided clinical expertise for development of the methodology. LW helped to write the manuscript. All authors reviewed the manuscript.

## Appendix

### 5.1 Pre-processing: dimensionality reduction

#### ‘Other’ category

Very infrequent categories are grouped into an ‘other’ bucket. A ‘very low’ threshold of 10 was chosen based on visual inspection of the frequency histograms, to represent the long tail of numerous categories into one category bucket.

Reductions Pathology results are reduced using Max and Min operators, as the extreme values are considered to be the most clinically significant. Demographic and procedure-related numerical fields are reduced with Mean, and if they are not expected to change, the first instance. One exception is BMI, where Max is considered more meaningful to the outcome. All the reductions are shown in Table 5.

**Table 5:** Reduction methods used when joining one-to-many relationships

| Field | Function |
| --- | --- |
| Age | Mean |
| BMI | Max |
| Height | Mean |
| Weight | Mean |
| Gender | First |
| Pathology tests (except EGFR) | First |
| Pathology EGFR | Sum |
| Medication | Sum |
| Length-of-stay (hours) | Mean |
| Unplanned 30-day readmission (Boolean) | Or |
| Discharge deceased (boolean) | Or |
| Discharge destination | First |

### 5.2 Hyperparameter tuning

We used Optuna for hyperparameter tuning, which utilises the TPESample algorithm and Hyperband pruner. We used a subset of the hyperparameters that made a difference empirically in preliminary runs with the baseline model–alpha, number of estimators, class balance and maximum depths.

## Footnotes

1 False Positive Rate is FP/(FP+TN). If FP is high, but TN is very large, the denominator remains high and the rate low.

